# Molecular Group and Correlation Guided Structural Learning for Multi-Phenotype Prediction

**DOI:** 10.1101/2023.12.26.23300559

**Authors:** Xueping Zhou, Manqi Cai, Molin Yue, Juan Celedón, Ying Ding, Wei Chen, Yanming Li

## Abstract

We propose a supervised learning algorithm to perform feature selection and outcome prediction for genomic data with multi-phenotypic responses. Our algorithm particularly incorporates the genome and/or phenotype grouping structures and phenotype correlation structures in feature selection, effect estimation, and outcome prediction under a penalized multi-response linear regression model. Extensive simulations demonstrate its superior performance over its competing methods. We apply the proposed algorithm to two omics studies. In the first study, we identified novel association signals between multivariate gene expressions and high-dimensional DNA methylation profiles, providing biological insights into how CpG sites regulate gene expressions. The second study is for cell type deconvolution. Using the proposed algorithm, we were able to achieve better cell type fraction predictions using high-dimensional gene expression data.

## Introduction

Rapid advances in modern bio-technologies have yielded massive omics data, including genetics, transcriptomics, proteomics, and epigenetics among many others. Such datasets are high-dimensional, where the number of features exceeds the sample size. In association studies, it is often that only a small proportion of the features are associated with the outcomes of interest. To identify the truly associated features among high-dimensional candidates, regularization methods are commonly employed to shrink the coefficients of noises to zeros. Another intrinsic characteristic of omics data is the genome hierarchy grouping structures. For example, single nucleotide polymorphisms (SNPs) can be grouped by their harboring genes, and genes can be grouped by functional pathways. Regularized regression methods with high-dimensional predictors that take into consideration of predictor grouping structures have been developed [30, 35], and they have been shown to provide better prediction performance compared to their counterparts without considering the predictor grouping structures.

Recent arising research interest has been focused on omics studies with multivariate responses. Two distinct approaches are usually adopted in multivariate response association studies. The first approach involves carrying out separate association studies on one response at a time. The second approach involves conducting one joint association study with all responses together. Statistical methods for the first approach have been widely studied. The top performers include, but not limited to, the least absolute shrinkage and selection operator (Lasso) [31], the elastic net [37], the group Lasso [35] and the sparse group Lasso [30]. One drawback of such an approach is that it treats response variables as independent and overlooks the correlation and grouping structures among the responses. Moreover, the first approach is typically followed by multiple comparisons for post-selection hypothesis testing, which can be stringent and result in excessive false negatives, particularly when the dimension of the responses is high.

The available methods for the second approach are still relatively limited. Li, Nan and Zhu [21] introduced the multivariate sparse group Lasso (MSGLasso), which performs signal selection between high-dimensional multivariate responses and multiple predictors while taking into account the grouping structures on both. MSGLasso outperforms the univariate response approaches in terms of outcome prediction and feature selection. However, MSGLasso does not explicitly consider the response covariance structure. Wilms and Croux [32] later proposed a multivariate group Lasso with covariance estimation that takes into account the covariance structure of the responses. However, it only considered the group-level sparsity, but not the within-group sparsity.

In this study, we propose an extension of MSGLasso – the **Brilliant**: Biological gRoup guIded muLtivariate muLtiple lIneAr regression with peNalizaTion. Same to MSGLasso, Brilliant utilizes the intrinsic genome hierarchy grouping structures in model fitting and predictions. It selects both the important group-level and therein the important individual-level association signals. Different to MSGLasso, Brilliant explicitly incorporates the covariance structure of multivariate responses. By doing so, the proposed Brilliant method improves the performance of feature selection, outcome prediction, and coefficient estimation significantly compared to MSGLasso and other univariate response approaches.

The rest of the paper is structured as follows. In section 2, we introduce the detailed algorithm of Brilliant. Section 3 uses simulation studies to demonstrate the performance of Brilliant. Section 4 presents two real data applications. A discussion is given in Section 5.

## Method

Let *n* denote the number of samples, consider multivariate linear regression model

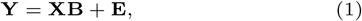

where **Y** = (**y**_1_, …, **y**_*q*_) ∈ ℝ^*n*×*q*^ is the response data matrix of *n* samples and *q* responses, and **X** = (**x**_1_, …, **x**_*p*_) ∈ ℝ^*n*×*p*^ is the predictor matrix of *n* samples and *p* predictors, **B** ∈ ℝ^*p*×*q*^ is the coefficient matrix, and **E** ∈ ℝ^*n*×*q*^ is the matrix of random errors. Assume that each row vector of **E** follows a multivariate normal distribution *N* (**0, Σ**) with a zero mean vector and a *q* ×*q* covariance matrix **Σ**.

Suppose **X** has *P* groups and **Y** has *Q* groups. They together introduce *P* ×*Q* intersection block groups [21] on the coefficient matrix **B**. Let 𝒢 be the set of all *P* × *Q* groups and **B**_*g*_ be a single block group in 𝒢, *g* ∈ 1, …J, *G* = *P* × *Q*. For a group **B**_*g*_, denote its *L*_2_ norm by 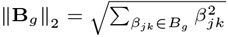.

Brilliant considers the following multi-responses optimization problem.

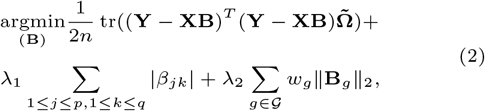

where 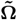 is a working version of the precision matrix **Ω** = **Σ**^−1^. When **Ω** is a diagonal matrix, Brilliant reduces to MSGLasso. Non-negative tuning parameters *λ*_1_ and *λ*_2_ are for the Lasso penalty and group Lasso penalty, respectively. The non-negative *w*_*g*_s are adaptive weights for each individual group. In this paper, we set 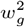 equal to total number of entries in the group *B*_*g*_.

The working precision matrix 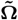 can be estimated from the data. For example, using the centralized responses, one can calculate 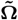 by directly invert the sample covariance matrix in a low dimensional setting or using graphical lasso [12] in a high-dimensional setting. In this paper, we propose to use a block diagonal working precision matrix with the diagonal blocks obtained by inverting the sample variance-covariance matrices of responses within each groups and the off-diagonal blocks set to be zero. We show that using a adequitely specified 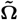, Brilliant can outperform MSGLasso.

Let 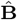 be the solution to (2) and *β*_*jk*_ be its entry on the *j*th row and *k*th column. For a given grouping structure 𝒢 on **B** and a given set of tuning parameters, an iterative algorithm adopted from the mixed coordinate descent algorithm [21] for coefficient estimation is summarized as in Algorithm 1. In the original mixed coordinate descent algorithm [21], the outcome covariance structure was not explicitly incorporated into the estimation. The Brilliant specifically incorporates the outcome covariance structure by integrating the user-specified working precision matrix into the mixed coordinating algorithm. In Algorithm 1, the estimates of *β*_*jk*_ by Brilliant depend on 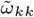 and *S*_*jk*_, where 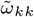 is the *kk*-the element (*k*-th diagonal element) of the working precision matrix 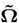 and 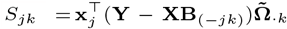 with **B**_(−*jk*)_ being the *jk*-th element of **B** replaced by zero and 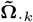 being the *k*th column vector of the working precision matrix. Incorporating the outcome covariance structure can be beneficial especially when the covariance structure of the outcomes is priory known or when the outcomes are highly correlated within or beteen groups. It is important to note that Brilliant incorporates both outcome correlations from within group and across groups, which is evidented in *S*_*jk*_. Please refer to Appendix 6 for a detailed proofs of the updating formules in algorithm 1.

To reduce false positives, we further threshold the estimated coefficient matrix 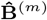 with a pre-defined non-negative threshold *b*_*thr*_. That is, 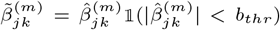, where 1 ≤ *j* ≤ *p*, 1 ≤ *k* ≤ *q*. Here *b*_*thr*_ is a hyper-parameter to be tuned from the data.

## Simulation studies

In our simulation studies, we compared the performance of Brilliant with that of the MSGLasso using the R package *MSGLasso* [21], and of the univariate method Lasso using the package *glmnet* [11]. The total number of predictors was set to 500. These predictors were separated into 10 non-overlapping groups with each group containing 50 predictors. Predictors within the same group were correlated with either a compound symmetry (CS) or a first-order auto-regression (AR1) structure with a correlation coefficient *ρ* = 0.5. Predictors from different groups were set to be independent. The predictor vector for each subject was independently generated from multivariate normal distributions with the specified correlation structures, marginal variances 1, and zero means. The total number of responses was set to be 100. Responses were separated into four non-overlapping groups and each group included 20 responses. For a given grouping structure G, we further assume the variables within a group are correlated, while variables from different groups are uncorrelated. That is equivalent to say that the true covariance matrix **Σ** assumes the form of a block diagonal matrix. The error vector for each subject was also independently generated from multivariate normal distributions with the same correlation structure as the predictor vector and a correlation coefficient *ρ* taking value in {0.1, 0.3, 0.5, 0.7, 0.9}. The covariance structures of each response group are specified in Table 1.

**Table 1.** Within-group covariance structures of **X** and **E** in each simulation setting.

| Setting | Covariance in $\mathbf{X}$ | Covariance in $\mathbf{E}$ |
| --- | --- | --- |
| 1 | CS, $\rho = 0.5$ | CS, $\rho = 0.1$ |
| 2 | | CS, $\rho = 0.3$ |
| 3 | | CS, $\rho = 0.5$ |
| 4 | | CS, $\rho = 0.7$ |
| 5 | | CS, $\rho = 0.9$ |
| 6 | AR1, $\rho = 0.5$ | AR1, $\rho = 0.1$ |
| 7 | | AR1, $\rho = 0.3$ |
| 8 | | AR1, $\rho = 0.5$ |
| 9 | | AR1, $\rho = 0.7$ |
| 10 | | AR1, $\rho = 0.9$ |

\* CS: compound symmetry.
\* AR1: first-order auto-correlation.
\* $\rho$ : correlation coefficient.

The coefficient matrix **B** was created to have a group structure with *P* × *Q* intersection groups. To achieve group-level sparsity of **B**, we randomly selected 1*/*5 of all groups to be non-zero groups, or groups with informative features. To attain within-group sparsity, we randomly selected 1*/*2 of the entries within a non-zero group and set to 0s. The other 1*/*2 non-zero values were draw from a uniform distribution U{[−3, 0) ∪ (0,, 3]}. **Y** was generated based on the multivariate multiple linear regression framework described in (1).

### Algorithm 1

Brilliant mixed coordinate descent algorithm

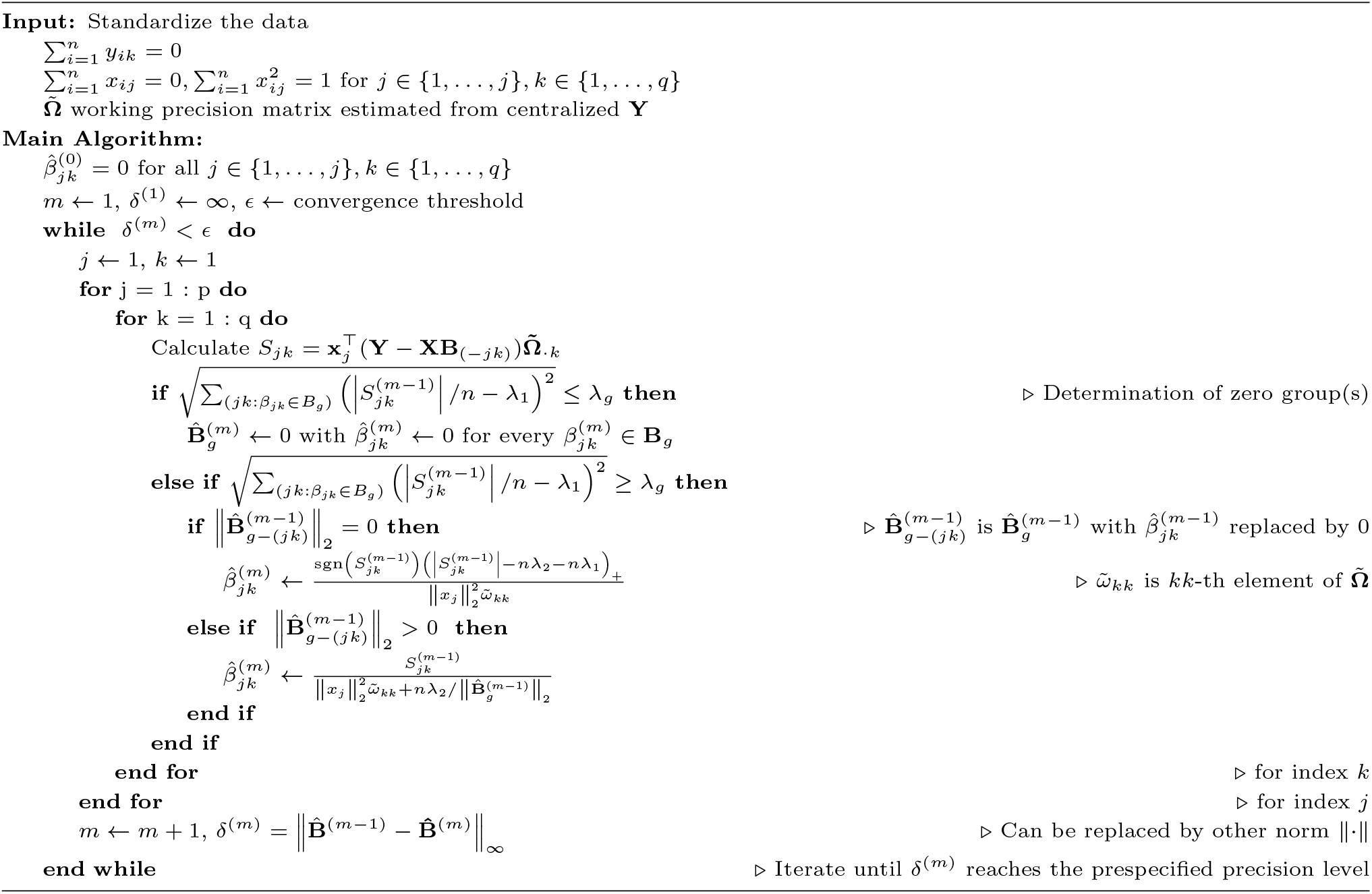

For each of the simulation scenarios, 100 experiments were independently replicated to evaluate the performance. Each experiment contained a training, a validation and a test dataset. We set the training sample size to 200, and the validation and test sample size to 100. We applied Brilliant on the training data using a block-diagonal working precision matrix with each diagonal block being the inverse of a within-group sample covariance matrix. A grid-search of selecting the optimal hyperparameters based on the prediction performance on the validation dataset was performed. The final prediction performance was evaluated on the test dataset.

The performance of outcome prediction, parameter estimation and feature selection was assessed. Outcome prediction was evaluated by average of the squared differences between the referenced cell fractions and their predicted values, or the mean square error (MSE), defined in (3), where *n* denotes the sample size in the test set. Feature selection and coefficient estimation were evaluated based on the estimated regression coefficient matrix 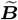. For coefficient estimation, mean absolute estimation error (MAEE) (4) was assessed. For feature selection, we calculated precision, recall and F1 score as formulated in (5)-(7), where |𝒮| is cardinality of a set 𝒮.

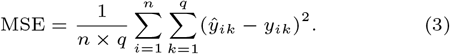

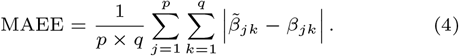

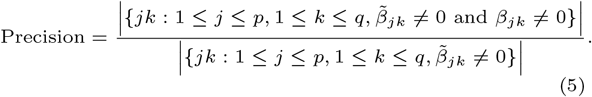

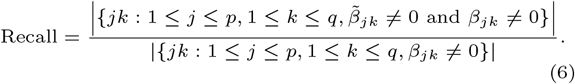

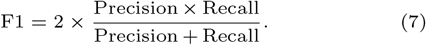

Figure 1 and Figure 7 summarize the performance of Brilliant, MSGLasso and Lasso when the covariance structure is CS or AR1, respectively. Brilliant outperforms MSGLasso and univariate lasso when the correlation *rho* within the same error group is 0.3 or greater. The other multivariate approach MSGLasso gives the second best prediction performance (Figure 1A) after Brilliant. This confirms that taking into account the multivariate response grouping structure can improve prediction compared to univariate response approach. It also shows that incoporating the responses’ covariances can further improve prediction. For coefficient estimation (Figure 1B), it shows that by explicitly incorporating the responses’ covariances, Brilliant obtains the lowest estimation error, especially when the responses are highly correlated. For feature selectio, as compared to MSGLasso and Lasso, Brilliant also gives higher precision, recall and F1 scores when response correlations range from moderate to high (Figure 1C-E). MSGLasso outperforms Brilliant when the correlation *rho* is low (*rho* = 0.1). This is due to the estimation error from calculting the precision matrix (inverting the working covariance matrix) in Brilliant.

**Fig. 1.**
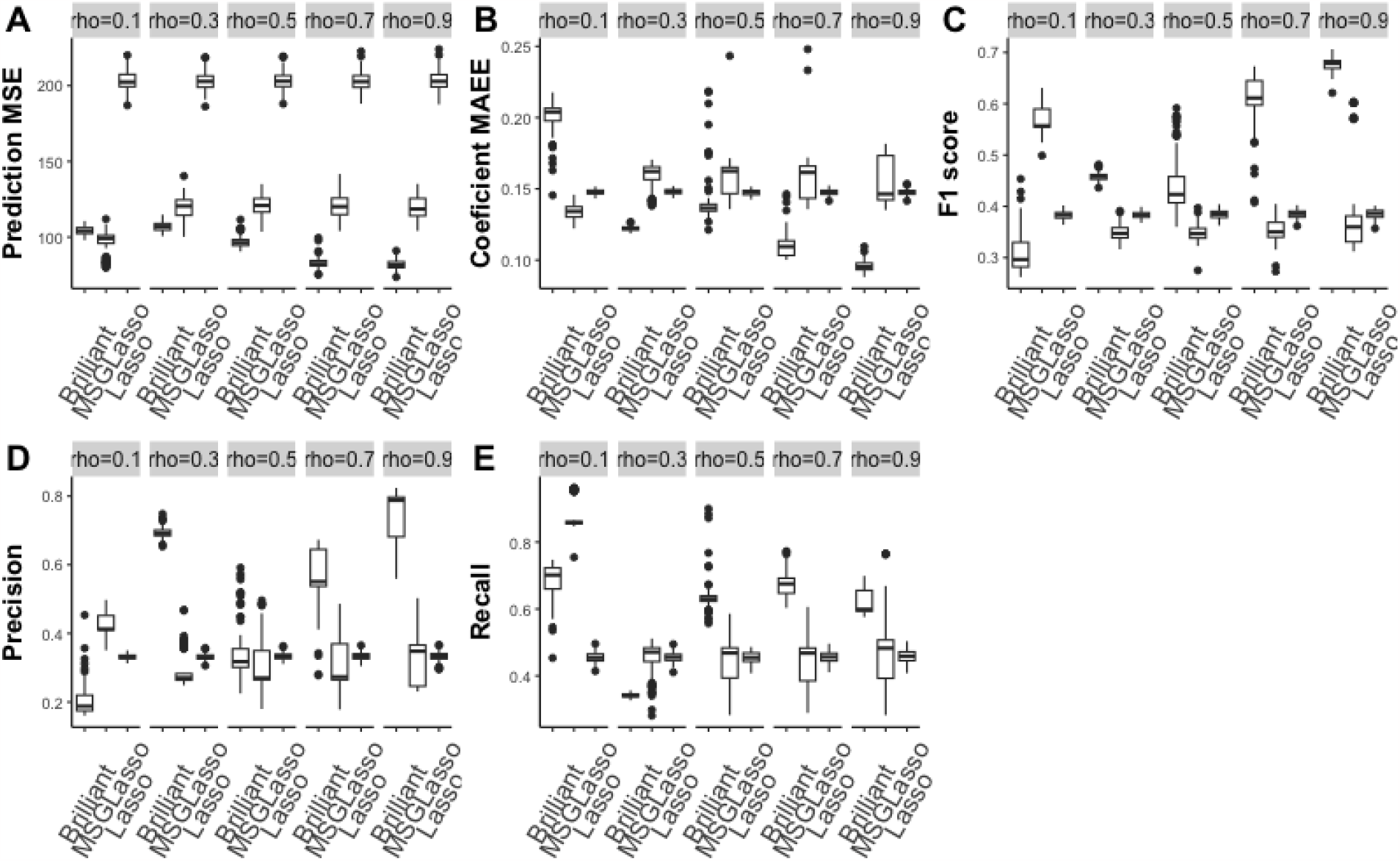
Simulation result of *p* = 500 with 10 **X** groups, *q* = 100 in 5 𝒴 groups. Covariance structure in each **X** group follows CS with correlation coefficient *ρ* = .5. Covariance structure in each 𝒴 group follows CS with *ρ* = 0.1, 0.3, 0.5, 0.7 or 0.9. Training sample size is 200 and test sample size is 100. **A**. Mean squared prediction error (MSE) with lower value indicating better prediction performance; **B**. Mean absolute estimation error of coefficient matrix (MAEE) with lower value suggesting better estimation; **C**. F1 score for feature selection; **D**. Precision for feature selection; **E**. Recall for feature selection; For **C**-**E**, a higher value indicates better feature selection performance.

## Real Data Application

We demonstrate the potential use and performance of Brilliant using the real data from our in-house multi-omics Epigenetic Variation and Childhood Asthma in Puerto Ricans (EVAPR) study [10] and the Framingham Heart Study (FHS) [24, 22]. In the EVAPR study, we collect phenotype, bulk RNA-sequencing (RNA-seq) gene expression and DNA methylation (DNAm) data from Puerto Rican children aged 9-20 years. Here, we focus on the 218 non-asthma controls, who have no missing data on all the predictors and outcomes. The FHS dataset is downloaded from dbGaP (phs000007.v32.p13), which contains blood microarray expression for 4,100 samples.

### Association analysis of DNA methylation and gene expression

In the first real data example, we applied Brilliant and its competing methods to the bulk RNA-seq gene expression and DNAm data from the nasal epithelium tissue samples of the 218 subjects to identify the susceptability CpG site associated with expressions of atopy-related genes.

The multivariate responses were expressions of 152 genes in four atopy-related biological pathways from the Gene Ontology Consortium [3]. Previous studies have shown that cytokine [26] and histone [1] play important roles in atopy. Here, we include 37 genes from the “positive regulation of cytokine production” pathway and 22 genes from the “histone methylation” pathway. We also include 55 genes in “energy derivation by oxidation of organic compounds” pathway since oxidative stress is directly related to atopic diseases [6]. Moreover, 39 genes from the “transport along mricrotuble” pathway were included due to their potential role in atopic diseases [15]. The average of within-pathway pairwise Pearson correlations was 0.715 (standard error= 0.168, details are given in Table 2 and Figure 2). DNAm information on 1,897 CpG sites around the 152 genes (within the region of 1, 000 base pairs up/down streams) were extracted. We further chosed the top 500 CpGs with the largest variances in their methylation levels (*M* values). The log-transformed gene expression TPM values and log-transformed methlylation *M* values were then used as the input responses and predictors for the Brilliant and MSGLasso. The groups of the gene responses were based on their belonging pathways. The groups of CpG sites were also based on the belonging pathways of their harboring or closest nearby genes. As a result, there were four gene groups and four CpG groups, which together yield 16 (= 4 × 4) interaction block groups on the underlying to be estimated regression coefficient matrix. The sizes of CpG groups vary from 91 to 143.

**Table 2.**
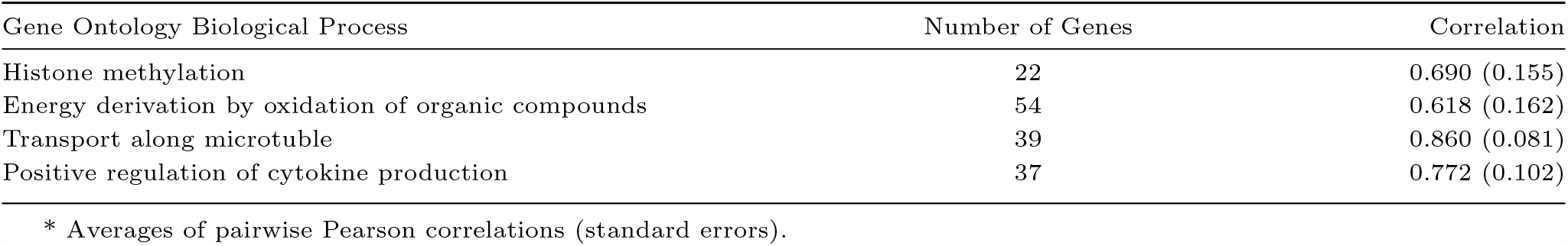
Average of pairwise Pearson correlations for genes within each outcome group.

**Fig. 2.**
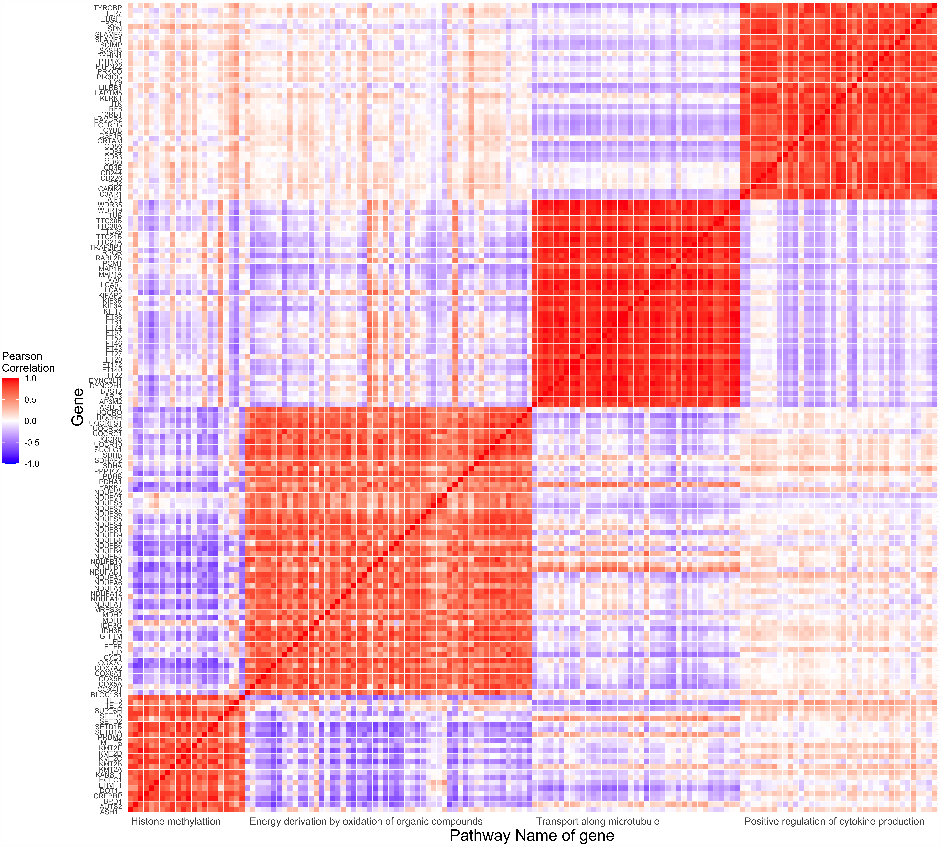
Pearson correlation heatmap of 152 genes. The vertical axis is indexed by gene names. The horizontal axis is indexed by the pathway names that the gene belongs to.

A three-fold nested cross-validation (CV) procedure was implemented for Brilliant and the competing methods. The inner CV was for selecting the optimal tuning parameters and the outer CV was for generating prediction results and avoiding overfitting. The Brilliant method outperforms both Lasso and MSGLasso in terms of giving the smallest predcition error per gene (Table 3). This is partially due to the the relatively high strength correlations between genes within each pathway group, and their un-ignorable roles in generating accurate prediction outcomes.

**Table 3.**
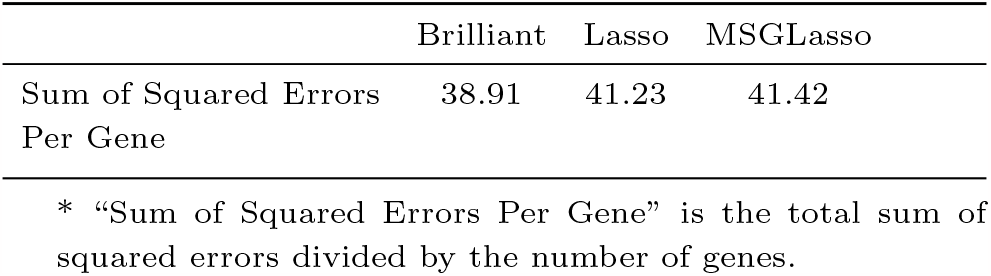
Prediction performance of Brilliant and its competing methods in the EVAPR Nasal gene expression and methylation association study.

Brilliant detected 7,600 potential associations between CpG methylations and gene expressions. MSGLasso and Lasso identified 2,575 and 1,728 associations, respectively. Among the 7,600 potential associations selected by Brilliant, 408 were also detected by both Lasso and MSGLasso (Figure 3). In the association block between the CpG group – “histone methylation” pathway and the gene group – “energy derivation by oxidation of organic compounds” pathway, Brilliant detected 96 association signals between DNAm and gene expression (Figure 9). There were 143 CpG sites in the “histone methylation” pathway and 54 genes in the “energy derivation by oxidation of organic compounds” pathway. The top association signals uniquely detected by Brilliant are listed in Table 4. Among them, majority of the associated CpG sites are located near genes known as key transcriptional and/or epigenetic regulators, with the associating genes being their targeted regulating genes. For instance, the association between *cg05608790* and *NDUFB9* was detected by Brilliant (*β*_*brilliant*_ = −0.0095). This reflects the regulation of *cg05608790* on its neighboring gene *AUTS2* (*β*_*brilliant*_ = −0.0150), which is an epigenetic regulator of a transcriptional network including *NDUFB9* [16]. Similarly, the association between *cg19832721* and *NDUFA4* (*β*_*brilliant*_ = −0.0104) reflects the regulational effects of *cg19832721* on its nearby gene *KANSL1* (*β*_*brilliant*_ = 0.0114), which is a key gene for producing the histone acetyltransferase 8 (KAT8, also known as MOF) regulatory nonspecific lethal complex. This complex is critical for acetylation of nucleosomal histone H4 at lysine 16, a unique histone mark controlling chromatin structure, and therefore involved in the regulation of transcription [28, 8, 29]. For this coefficient block, Lasso detects an additional 237 potential CpG-Gene associations. In contrast, MSGLasso detected only 7 association signals within this block.

**Table 4.** Top association signals between DNA methylation and gene expression uniquely selected by Brilliant.

| CpG site | Gene | Gene Chr | CpG Position | CpG Nearby Gene | $\hat{\beta}$ | Group |
| --- | --- | --- | --- | --- | --- | --- |
| cg04494844 | AUTS2 | 7 | 7:70257926 | AUTS2 | 0.0182 | I-1 |
| cg04494844 | NDUFV2 | 18 | 7:70257926 | AUTS2 | 0.0138 | I-2 |
| cg05608790 | AUTS2 | 7 | 7:69427388 | AUTS2 | -0.0150 | I-1 |
| cg05608790 | NDUFB9 | 8 | 7:69427388 | AUTS2 | -0.0095 | I-2 |
| cg05608790 | PDHB | 3 | 7:69427388 | AUTS2 | -0.0111 | I-2 |
| cg20134287 | AUTS2 | 7 | 7:70061455 | AUTS2 | -0.0208 | I-1 |
| cg20134287 | NDUFS5 | 1 | 7:70061455 | AUTS2 | -0.0192 | I-2 |
| cg20134287 | PDHA1 | X | 7:70061455 | AUTS2 | 0.0152 | I-2 |
| cg19832721 | KANSL1 | 17 | 17:44249866 | KANSL1 | 0.0114 | I-1 |
| cg19832721 | NDUFA4 | 7 | 17:44249866 | KANSL1 | -0.0104 | I-2 |
| cg19832721 | NDUFB9 | 8 | 17:44249866 | KANSL1 | -0.0122 | I-2 |
| cg18829443 | TET3 | 2 | 2:74227916 | TET3 | -0.0171 | I-1 |
| cg18829443 | NDUFB4 | 3 | 2:74227916 | TET3 | 0.0181 | I-2 |

\* $\hat{\beta}$ is the estimated regression coefficient. “Chr” stands for chromosome.
\* CpG positions were mapped to hg19 reference genome.
\* Group I denotes the genes in the the “histone methylation” pathway. Group 1 denotes the CpG sites within genes in “histone methylation” pathway. Group 2 is the CpG sites within genes in “energy derivation by oxidation of organic compounds” pathway. Group I-1 is the coefficient block corresponds to associations between genes in the “Histone methylation” pathways and CpG sites within genes in the same pathway. Group I-2 is the coefficient block corresponds to associations between genes in the “histone methylation” pathways and CpG sites within genes in the “energy derivation by oxidation of organic compounds” pathway.

**Fig. 3.**
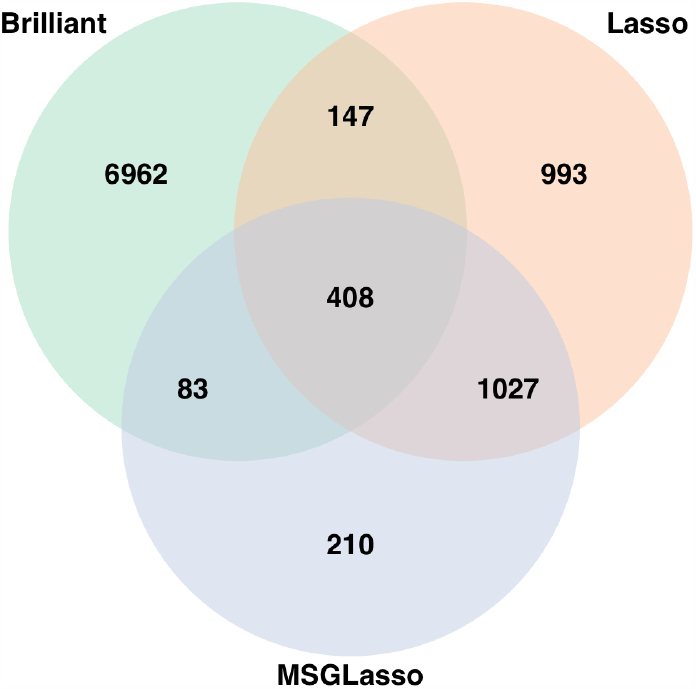
Venn diagram of numbers of association signals detected by Brilliant and the competing methods in EVAPR study.

### Cell-type deconvolution for bulk gene expression in blood cell sample

Tissue-level omics studies are based on the average effects across heterogeneous collection of cells, such as studies on peripheral blood include signals from all underlying cell types. Therefore, the tissue-level association studies are prone to be confounded by the variation in cell composition across samples [19]. To eliminate such confounding, different algorithms of cell-type deconvolution have been proposed to infer the fractions of cell types in bulk gene expression data collected from tissue samples [4]. Among available methods, the reference-based cellular deconvolution are believed to be the most accurate and reliable approach [4, 17]

In the second real data application, we used both the EVAPR and FHS datasets to predict cell type proportions for blood neutrophil (Neutro), monocyte (Mono), lymphocyte (Lymph) and eosinophil (Eosino) using the bulk gene expression data from blood cell samples. The multivariate responses for both datasets were the fractions of the four cell types for each subject, the values of which were measured through complete blood cell counts [20]. We put all four cell type fractions into one response group for both datasets. Predictors in EVAPR dataset were expression levels of the signature genes of each cell type based on the widely used leukocyte gene signature matrix reference (LM22) [25]. Those genes were put into four predictor groups based on their reference cell type groups. More specifically, there were 64 signature genes in neutrophil group, 53 in lymphocyte group, 40 in monocyte group, and 39 genes in eosinophil group. The transcripts per million (TPM) counts of predictors were first undertaken a log transformation before applying the Brilliant algorithm. Predictors in FHS dataset contains blood microarray gene expressions from 4,100 samples Those genes were also grouped into four groups based on the leukocyte signature genes available in the dataset (Details in Appendix 9). The microarray gene expression data were also log transformed.

Prediction performance of Brilliant were evaluated within each and also across the two datasets. For internal prediction within each datasets, a three-fold nested cross-validation was perfromed on each dataset to select the optimal hyperparameters and avoid overfitting. For cross dataset prediction, FHS data were used as the training data and EVAPR data were used as the test data. A three-fold cross-validation was performed on the FHS data to select the optimal hyperparameters. The prediction results are shown in Figure 4, where the scatter plots between the predicted and referenced cell fractions and their Pearson correlations were presented. The correlations range from 0.692 to 0.896.

**Fig. 4.**
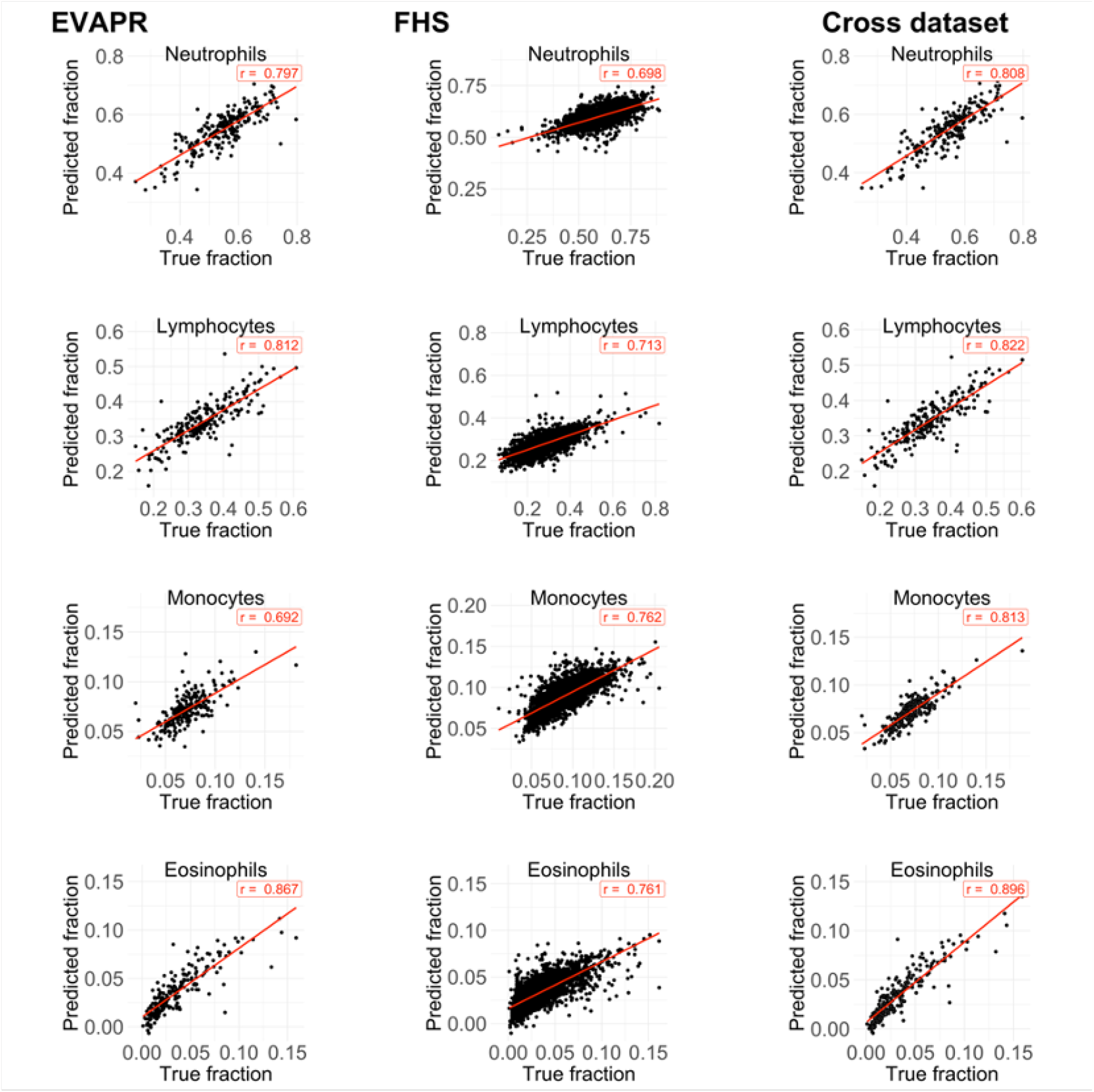
Scatter plots of predicted v.s. referenced cell type fractions. The left panel plots are from CV using the EVAPR data only. The middle panel are from CV using the FHS data only. The right panel are from using EVAPR data as the test set while using the FHS data as the training set.

**Fig. 5.**
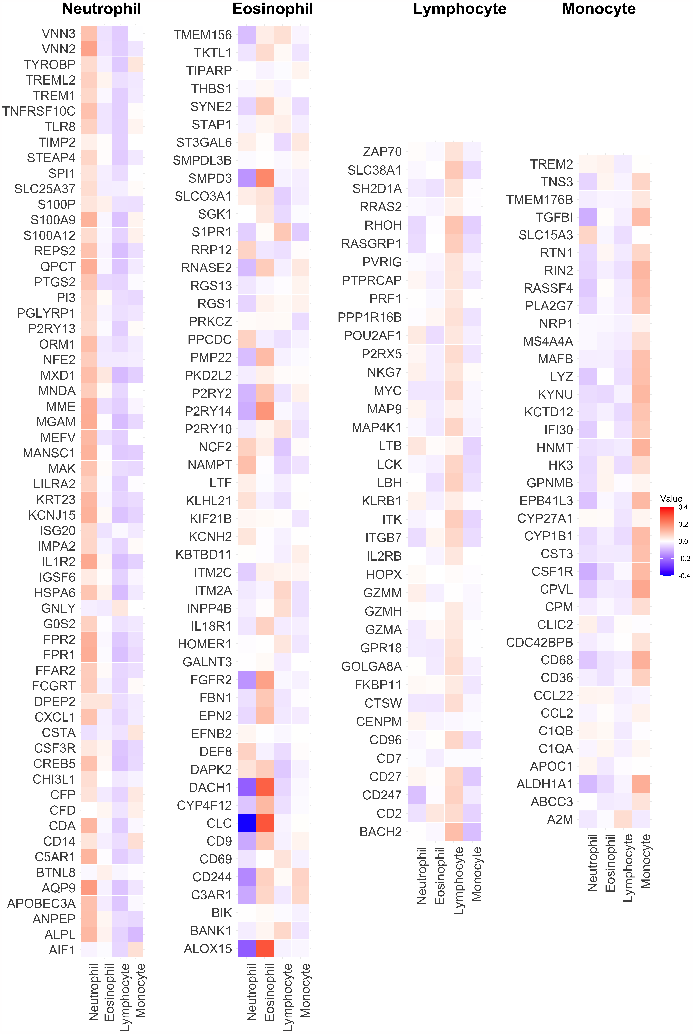
Heatmap of coefficient matrix estimated by Brilliant using the FHS data. The matrix has been divided into four blocks based on the signature genes of each cell type. The four blocks contain the signature genes of Neutrophil, Eosinophil, Lymphocyte, and Monocyte cells, respectively.

The predicted cell type fractions from Brilliant were compared with other commonly used reference-based cell-type deconvolution methods, as in the study by Cai et al. [7], including CIBERSORT/CIBERSORTx [25], DeconRNASeq [13], DCQ [2], DeCompress [5], DSA [36], dtangle [17], EnsDeconv (Ensemble Deconvolution) [7], GEDIT (Gene Expression Deconvolution Interactive Tool) [23], EPIC (Estimating the Proportions of Immune and Cancer cells) [27], FARDEEP (Fast And Robust DEconvolution of Expression Profiles) [14], hspe (hybrid-scale proportions estimation) [18], ICeDT (Immune Cell Deconvolution in Tumor tissues) [33], and SCDC methods including non-negative least squares (SCDC NNLS) and inverse sum of squares (SCDC ISSE) [9]. All the results were based on the LM22 reference. The FHS dataset was used as the training data, and the EVARP dataset was used as the test data. The comparison results are given in Figure 6, where the Spearmans’s rank correlation, a convention performance evaluation metric in the cell-type deconvolution field, between the predicted and referenced cell fractions for each of the aforementioned methods. Although not designed as a cell-type deconvolution algorithm, Brilliant achieves the highest average Spearman’s correlation (the vertical bars in Figure 6) across the four cell types. We also calculated the mean square of errors (MSE) between the predicted and referenced cell fractions for each of the methods (Figure 10 in the Appendix). Brilliant gave the smallest average MSE across cell types among all methods.

**Fig. 6.**
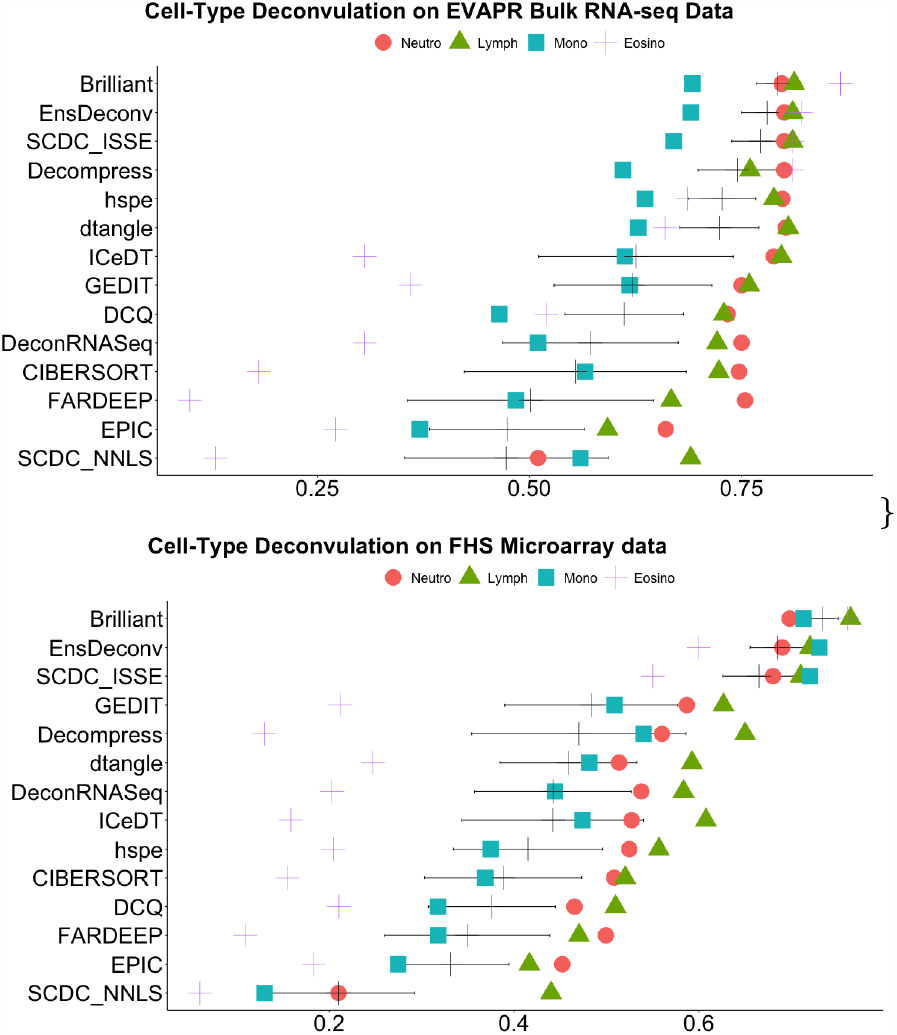
Comparison of cell-type fraction prediction using Brilliant and different deconvolution methods. Neutro: neutrophil; Mono: monocyte; Lymph: lymphocyte; Eosino: eosinophil. The black vertical bar on each row shows the mean of Spearman’s correlations, and the horizontal line presents the range of one standard error below/above the mean.

**Fig. 7.**
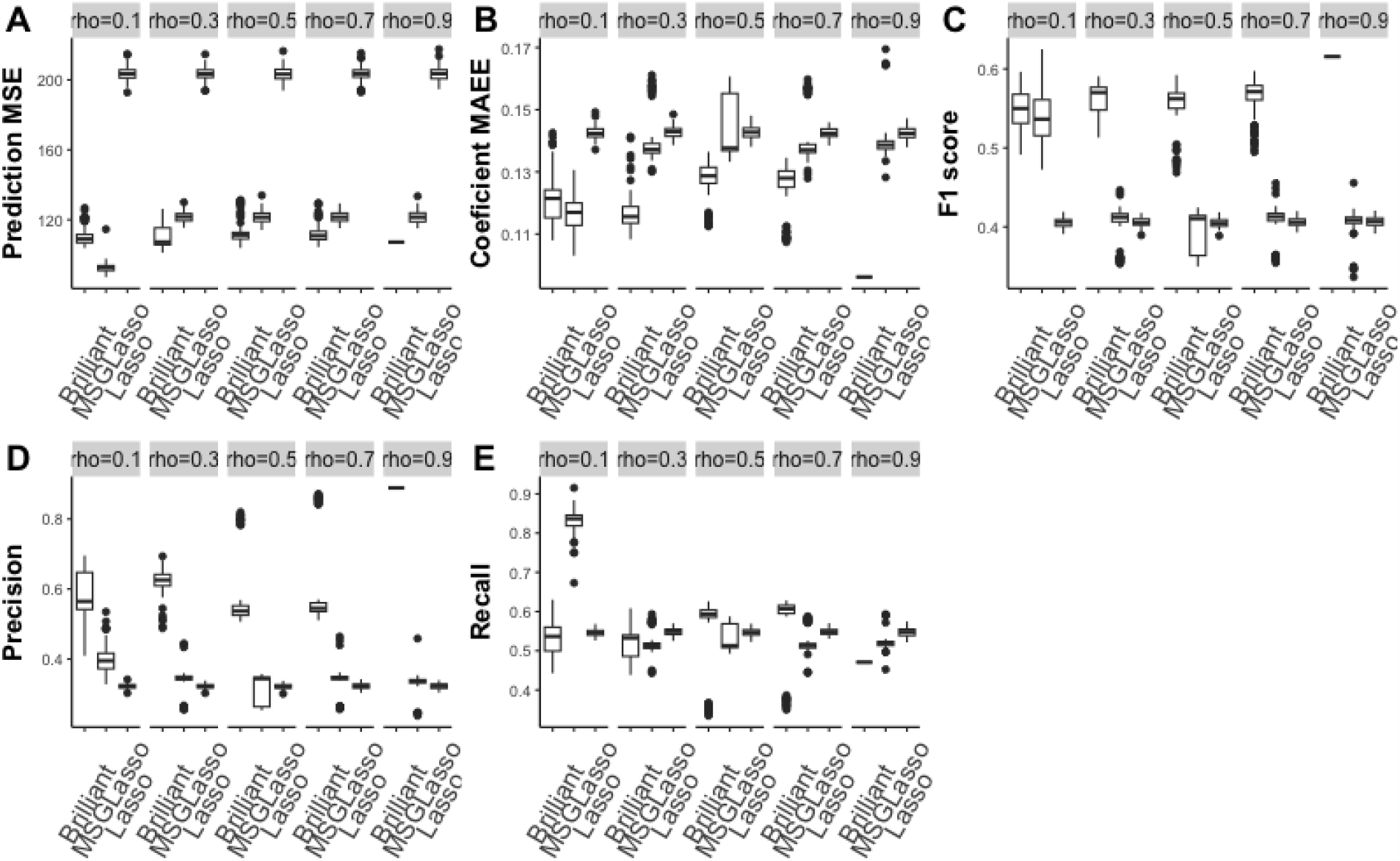
Simulation result of *P* = 500 in 10 groups, *Q* = 100 in 5 groups, covariance structure of each predictor group is first-order auto-regressive (AR1) with associated correlation coefficient *ρ* = .5, covariance structure of each error group is CS with*ρ* = .1, 0.3 .5, .7, or .9. Training sample size is 200. **A**. Mean squared error (MSE) metric of prediction error, and lower value means better prediction performance; **B**. Mean absolute estimation error (MAEE) of coefficient matrix with lower value suggesting better estimation **C**; F1 score for feature selection; **D**. Precision for feature selection. **E**. Recall for feature selection; For **C**-**E** higher value indicates better feature selection performance.

**Fig. 8.**
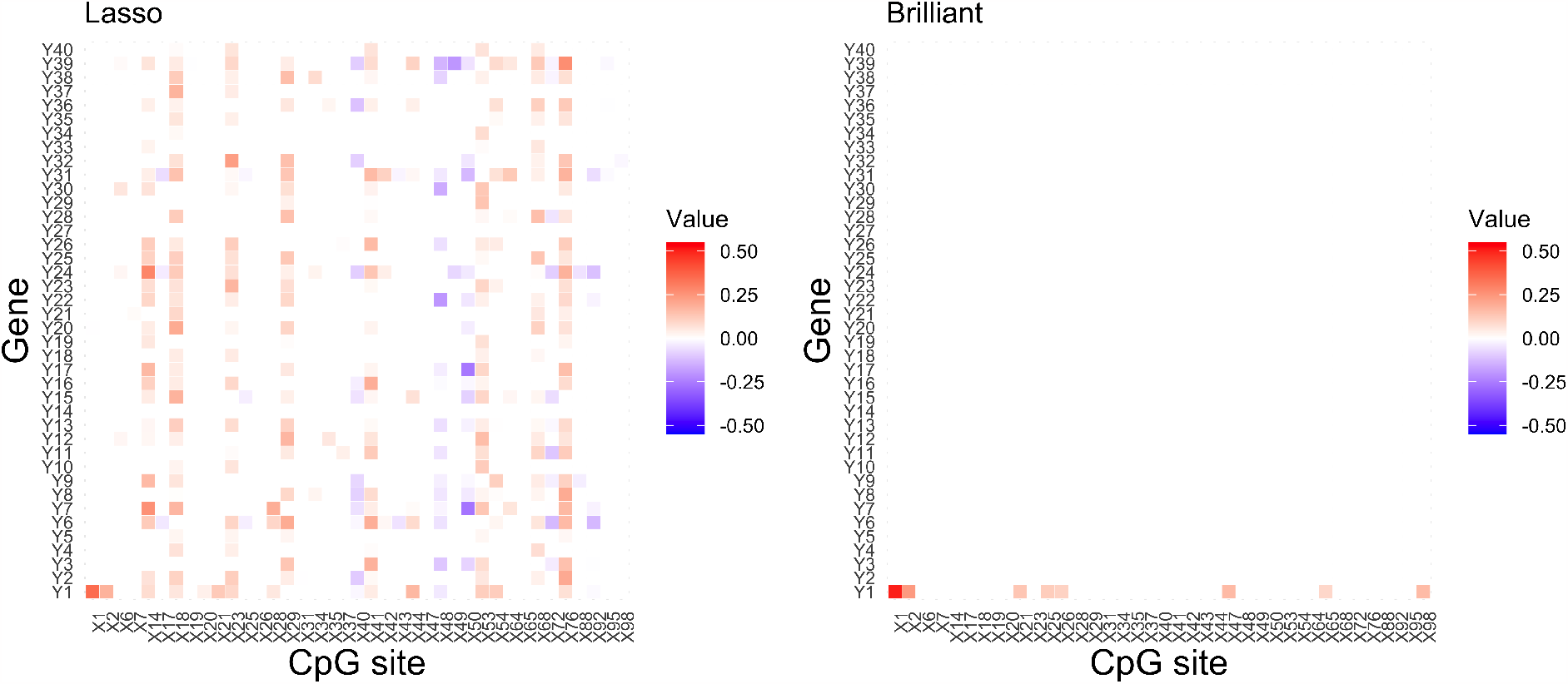
Heatmaps of estimated coefficient to demonstrate Lasso select more false positives when the multivariate outcomes are correlated. Note: Predictors without any nonzero estimated coefficients by both Lasso and Brilliant are omitted in the graph.

**Fig. 9.**
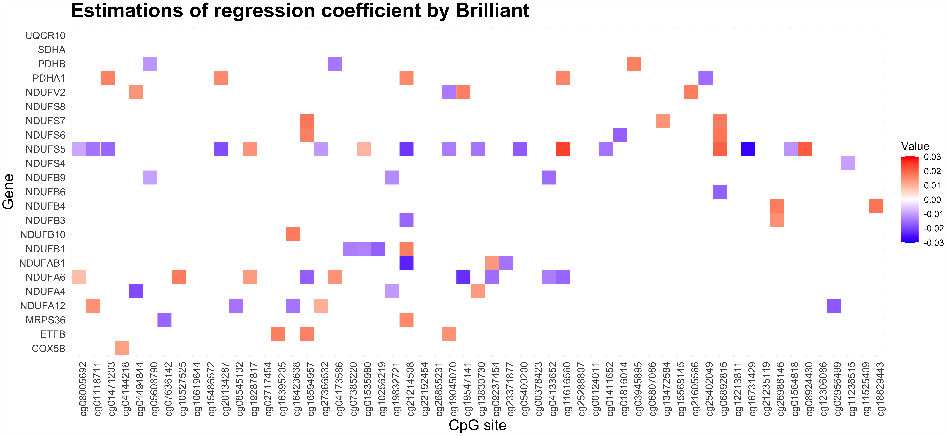
Estimations of one regression coefficient block by Brilliant. Note: This example coefficient block corresponds to the associations between the 143 Cpg sites near genes from the “histone methylation” pathway and the 54 genes from the “energy derivation by oxidation of organic compounds” pathway. Only CpG sites and genes with at least one non-zero coefficient estimate are visualized in the graph.

**Fig. 10.**
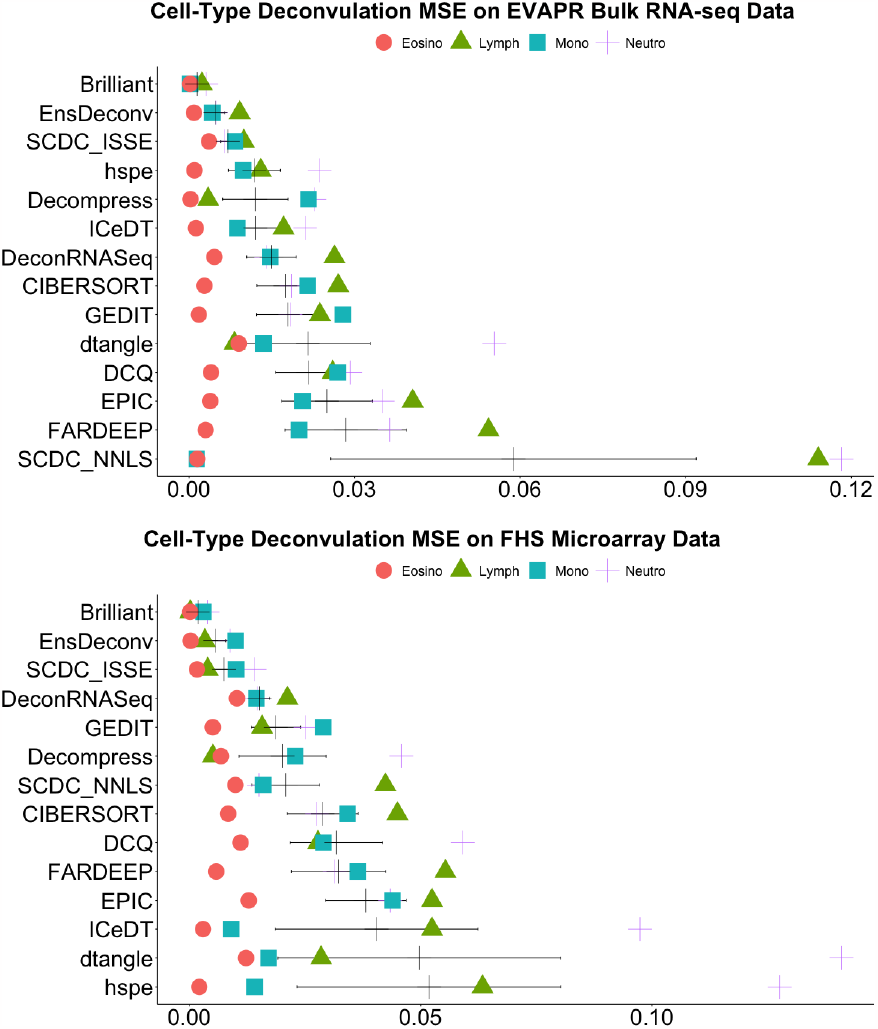
Comparison of cell-type fraction prediction mean squared error (MSE) using Brilliant and different deconvolution methods on white blood bulk RNA-sequencing data in the EVA-PR study and blood microarry data in the FHS study. Neutro is neutrophil; Mono is monocyte; Lymph is lymphocyte; and Eosino is eosinophil. Dot denotes the MSEs between the predicted fraction and truth for one specific cell type. The black vertical bar shows the mean of cell-type specific MSEs, and the horizontal line presents mean *±* standard error of the mean.

The feature selection performance of Brilliant was evaluated by comparing the estimated coefficient matrix in the final model (built on FHS data) with the LM22 reference gene matrix. The heatmap in Figure 5 displays the coefficient matrix estimated by Brilliant. The matrix has been divided into four sub-matrices based on the signature genes of each cell type. As anticipated, for each cell type, the signature genes in the LM22 reference have the highest estimate magnitudes in their corresponding cell type categories (indicated by darker red color in the corresponding column). Among the 188 genes selected in the final model, Brilliant correctly identified cell types for 144 signature genes. In other words, the highest proportioned cell type in the LM22 matrix is also estimated by Brilliant to have the highest coefficient magnitude across four cell types for these 144 genes. For another 20 genes, the cell type with the second-largest estimated coefficient magnitude is the highest proportioned cell type in the LM22 reference. These demonstrate that Brilliant is capable of identifying the most informative features associated with the outcomes of interest.

## Conclusion

In this paper, we proposed an analytical tool – Brilliant for variable selection and outcome prediction on multivariate response and high-dimensional predictor data. Brilliant not only utilizes the intrinsic grouping structures of both predictors and responses and performs both group-level and within group-level variable selection, but also explicitly incorporates the covariance structure of the multivariate responses. Brilliant significantly improves the coefficient estimation accuracy, feature selection and outcome prediction performance compared to the competing methods. Brilliant does not require iteratively estimating the precision matrix as in [34]. It only requires a one-step estimation of a working covariance structure according to the user specified grouping structure on the responses.

We applied Brilliant to a methylation–to–gene expression association study and a study using gene expressions to predict cell type fractions. Brilliant can be easily extended to solve many other multivariate-response and multiple-predictor problems, including but not limited to single cell multiomics and expression quantitative trait loci (eQTL) studies, which are beyond the scope of this paper. We will explore its wide applications in the future.

## Data Availability

All data used in the present study are available upon reasonable request to the authors.

## Appendix A Proofs of technical results

### Brilliant objective function

Assume **Y**|**X** ∼ *N* (**XB, Σ**), the log-likelihood of data can be written as follows.

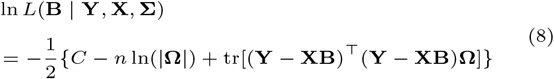

Given the working precision matrix 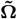, the penalized log likelihood can be written as below.

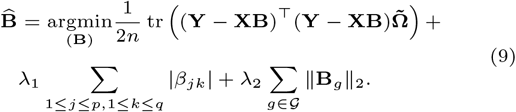

### Derivative of Brilliant objective function

The objective function consists of three terms. We take the derivative of each term with respect to *β*_*jk*_ separately. Then, we will combine the gradient or sub-gradient together.

#### Take derivative of the first term with respect to β_jk_

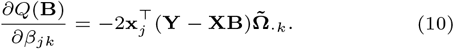

Let **B** = **B**_(−*jk*)_ + **B**_(*jk*)_, where **B**_(−*jk*)_ being the *jk*-th entry of **B** replaced by zero and **B**_(*jk*)_ being all but the *jk*-th entry of **B** replaced by zeros. Define *S*_*jk*_ as shown below in equation

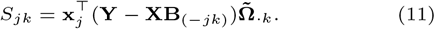

We can re-write equation (10) as

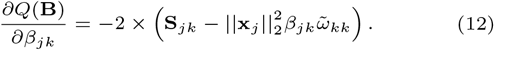

#### Take derivative of the second and third term with respect to β_jk_

For a coordinate *β*_*jk*_, when *L*(**B**) is differentiable at *β*_*jk*_,

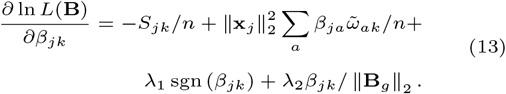

For general case, the derivative of the second term or |·| penalty can be written as follows.

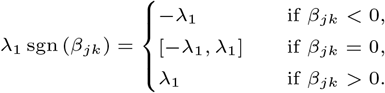

For general case, the derivative of the third term in the objective function or the derivative of ∥ · ∥_2_ penalty are shown below.

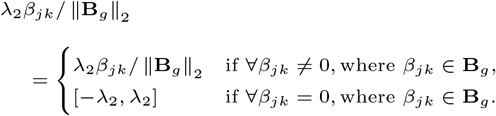

#### Combine the derivative of the three terms

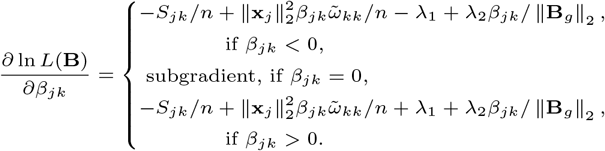

### Solution to the Brilliant objective function

Set derivative to 0 and solve for *β*_*jk*_ with the corresponding constraint, we can get

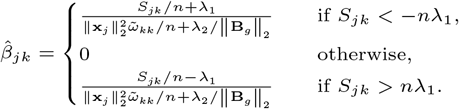

Or, in one unified form:

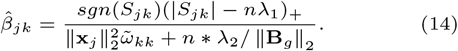

For those who wish to delve deeper into the topic, we have included a comprehensive explanation below.

#### Details when β_ij_ < 0

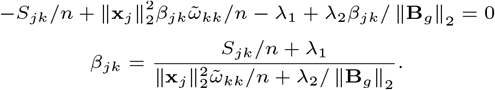

With the constraint *β_jk_ <* 0, or 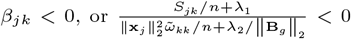, we have

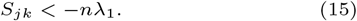

#### Details when β_ij_ > 0

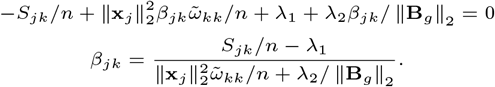

With the constraint *β_jk_ >* 0, or 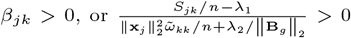, we can get

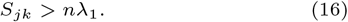

#### Details on β_jk_ = 0

We will see different scenarios involving *β*_*ij*_ = 0.

S1. The group containing *β*_*jk*_ is not zero groups. This suggests that the Lasso (*L*_1_) | · | penalty is not differentiable, but the ∥ · ∥ penalty is differentiable for all groups containing *β*_*jk*_. Set the derivative to 0, we get

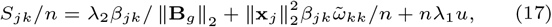

where |*u*| *<* 1. When *u* ≥ 0, *S*_*jk*_ ≥ *nλ*_1_. When *u <* 0, *S*_*jk*_ *<* −*nλ*_1_. So the solution is the same as that in the above session for *β*_*jk*_ ≠ 0) or Equation (14).

S2. The group containing *β*_*jk*_ is a zero group. Let 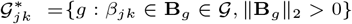. Let **B**_*g*0_ be the zero group containing *β*_*jk*_. Let |*u*| *<* 1, and ∥**v**∥_2_ *<* 1. Set the derivative to 0, we have

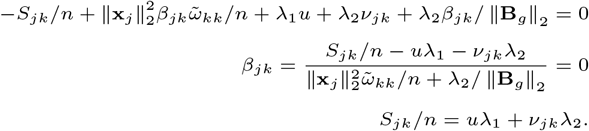

For each (*jk* : *β*_*jk*_ ∈ *B*_*gi*_),

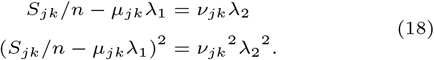

We can see 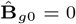 if

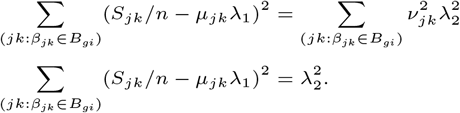

Discuss on a case by case basis,

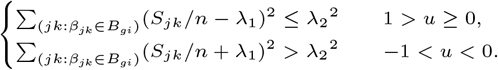

We can get that 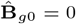

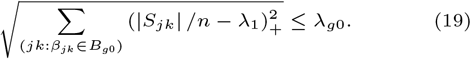

## Appendix B: Simulation Result

For *p* = 500 and *q* = 100, the simulation results under compound symmetry (CS) covariance structure (setting 1-5 in Table 1) are presented in the main article. For results under first order autoregressive (AR1) covariance structure (setting 6-10 Table 1), we observe very similar performance of Brilliant as in CS settings. More specifically, Brilliant achieves improved prediction and coefficient estimation compared to MSGLasso and univariate Lasso. Brilliant also shows significant improved feature selection performance, especially in F1 score and precision.

## Appendix C: Further explanation of Lasso feature selection performance

We use a simply simulation study to demonstrate that Lasso is susceptible to selecting more FP signals when there exist correlated outcomes. In this example, we simulated a group of 100 predictors and 40 outcomes. The predictors were generated using the multivariate normal distribution with covariance matrix following compound symmetry (cs) structure with *ρ* = 0.2. The outcomes were simulated from the multivariate normal distribution with covariance matrix following compound symmetry structure and *rho* = 0.9. Then we replaced *Y* 1 with *Y* 1 + *X*1 + *X*2. There were 25 observations in the training dataset. In this case, although both Lasso and Brilliant select the true signals, namely *Y* 1 − *X*1 and *Y* 2 − *X*2. Lasso identify an additional 331 false positives (FPs), while Brilliant only have 6 more FPs (Figure 8).

## Appendix D: Details on cell-type deconvolution for blood cell samples

In the LM22 reference matrix, all entries are positives with higher value indicating higher expression level of the gene in the corresponding cell type. For each gene, we set it as the signature gene for the cell type with the largest value in the reference matrix. The number of available signature genes in the EVAPR and FHS dataset are summarized in Table 5. Table 6 summarizes the top performers of cell type deconvolution for the EVAPR dataset.

**Table 5.** Signature genes available in EVAPR and FHS dataset.

| Dataset | Total number of genes | Neutrophil | Lymphocyte | Monocyte | Eosinophil |
| --- | --- | --- | --- | --- | --- |
| EVAPR | 200 | 64 | 53 | 40 | 39 |
| FHS | 196 | 61 | 40 | 40 | 55 |
| Overlap | 188 | 59 | 51 | 37 | 37 |

\* The values denote the number of signature genes.
\* The overlapping genes in the two datasets are used to build the final model using the FHS data, and predict the cell type fractions for the EVAPR data.

**Table 6.** Top performers of cell type deconvolution for EVAPR dataset.

| Method | Neutrophil | Lymphocyte | Monocyte | Eosinophil | Mean |
| --- | --- | --- | --- | --- | --- |
| Brilliant | 0.80 | 0.81 | 0.69 | 0.87 | 0.79 |
| Brilliant (Cross dataset) | 0.77 | 0.79 | 0.71 | 0.89 | 0.79 |
| EnsDeconv | 0.80 | 0.81 | 0.69 | 0.82 | 0.78 |

\* The values under each cell type are cell type specific Spearman’s correlation. Mean denotes the average of the four cell-type specific Spearman’s correlation.
\* Brilliant (Cross dataset) shows the performance of the final model (built using the FHS data) on predicting the cell type fractions for the EVAPR data.

## Competing interests

The authors declare no competing interest is declared. This study was partially supported by National Science Foundation (award number 2225775).

## Author contributions statement

X.Z., Y.L. and W.C. conceived and designed the study. M.Y. and J.C. was responsible for data management. X.Z. and M.C. were responsible for data cleaning and analysis. X.Z., Y.D., Y.L. and W.C. were responsible for interpretation. X.Z. and Y.L. wrote the paper. All authors read and approved the final article.

## Acknowledgments

The authors thank all the participants and their families in the Epigenetic Variation and Childhood Asthma in Puerto Ricans study and Framingham Heart Study for enabling this research and further our understanding of childhood asthma and allergy by their participation in the studies. We appreciate the researchers of the Framingham Heart Study for making the data available on dbGAP. This research is supported in part by the University of Pittsburgh Center for Research Computing through the resources provided. Specifically, this work used the HTC cluster, which is supported by NIH award number S10OD028483, and the H2P cluster, which is supported by NSF award number OAC-2117681.

## Notes

### Competing Interest Statement

The authors have declared no competing interest.

### Author Declarations

1. Epigenetic Variation and Childhood Asthma in Puerto Ricans (EVAPR) study (methylation, gene expression, and cell fraction data). 2. Framingham Heart(FHS) Study (gene expression and cell fraction data).

